# PHOENIX-f: A Randomised Controlled Feasibility Study of One-Stop Osteoporosis Screening among Patients Undergoing Routine Computed Tomography using FRAX and CliniQCT Bone Density Measurement and Vertebral Fracture Detection

**DOI:** 10.1101/2025.07.17.25331283

**Authors:** Daniel DG Chappell, Jane Fleming, Judith Brown, Rahul Shah, Emma M Clark, Helen Risebro, Lee Shepstone, Thomas D Turmezei, Adam P Wagner, Karen Willoughby, Stephen K Kaptoge, Kenneth E. S. Poole

## Abstract

*Objectives:* The PHOENIX-f (Picking up Hidden Osteoporosis Effectively during Normal CT Imaging without additional X-rays) study examined the feasibility of: i) screening for osteoporosis in patients undergoing computed tomography (CT) scans relating to other conditions using a one-stop service across multiple hospitals and; ii) a full trial to evaluate its effectiveness and cost-effectiveness. CT scan images of older patients at higher risk of fractures were opportunistically ‘re-used’ - through digital screening - to improve identification, treatment and fracture prevention.

*Design:* Randomised controlled feasibility study to inform the design of a future definitive trial by determining ability to randomise and collect follow-up measures and estimating this population’s osteoporosis and vertebral fracture prevalence, and treatment rates.

*Setting, Participants and Intervention:* In one teaching hospital and four regional UK hospital radiology waiting areas, patients undergoing abdomen ± pelvis CT scans (women aged ≥65, men aged ≥75) were offered ‘FRAX’ fracture risk questionnaires with embedded consent forms. Consenting patients identified as having moderate/high 10-year fracture risk based on initial screening (FRAX red/amber zones) were randomised (1:1:1) to: 1) CliniQCT One-Stop osteoporosis screening pathway [vertebral fracture assessment and hip/spine BMD measurement by applying Mindways QCT and SlicePick-MT software to CT scans, plus algorithm-based recommendations sent by electronic letter to general practitioners (GPs)]; 2) FRAX answers sent to GPs; 3) usual care (GP informed of participation).

*Main outcome measurements:* Co-primary feasibility endpoints were ability to randomise 375 patients within 10 months and retain 75% of survivors for a 1-year bone health outcome questionnaire.

*Results:* From 1,828 eligible for invitation, 595 participants consented; low FRAX scores excluded 213 patients. Outcomes achieved included randomising 382 patients within 10 months and 284/329 survivors (86%) completing 1-year follow-up. Of 356 analysable CT scans osteoporosis was diagnosed in 42%. 46/258 (18%) whole thoraco-lumbar scans showed vertebral fractures. At follow-up 36%, 26% and 8% of patients needing osteoporosis treatment in the screened, FRAX-only and usual care groups respectively reported taking treatment. Collecting data needed for a future economic evaluation to determine cost-effectiveness appeared feasible.

*Conclusions:* Both the CliniQCT screening pathway, and a trial to determine its effectiveness and cost-effectiveness, were shown to be feasible. Even within this small sample, we found relatively high rates of osteoporosis and a signal suggesting that CliniQCT could facilitate its treatment, warranting a full trial evaluation.

*Trial registration:* ISCRTN: 14722819 DOI https://doi.org/10.1186/ISRCTN14722819 Secondary identifying numbers: PB-PG-0816-2007; CPMS: 41112

**Summary box:** **What is already know about this topic**

- Vertebral fractures due to osteoporosis cause severe pain, increased disability and poor quality of life, are associated with higher subsequent fracture rates and mortality, yet are often not diagnosed.
- Bone preserving treatments can reduce osteoporotic fractures, including vertebral fractures.
- CT scans can be used to diagnose osteoporosis and vertebral fractures without additional radiation exposure but are not routinely used in this way

**What this study adds**

- Opportunistic bone density screening using CliniQCT and vertebral fracture assessment is feasible in older adults identified by FRAX as at moderate/high fracture risk.
- Osteoporosis and vertebral fracture prevalence is high in this population: in our study sample, 42% of QCT analysed scans showed osteoporosis and 18% of full thoraco-lumbar spine CTs showed vertebral fractures. Of all participants with vertebral fractures, 42% had multiple vertebral fractures.
- In this feasibility study initiation of recommended osteoporosis treatment by a year later was over 4 times higher for patients on the screening pathway than for patients receiving usual care (36% vs 8%).

**How this study might affect research, practice or policy**

- Embedding CT-based osteoporosis detection into routine imaging could markedly improve diagnosis and treatment uptake in high-risk patients and warrants full effectiveness and cost-effectiveness evaluation.

## Introduction

Osteoporosis is characterized by thin, porous bones and increased risk of fractures. In England and Wales, approximately 180,000 individuals are affected by osteoporosis-related fractures annually (1). Systematic reviews estimate that fragility fractures cost the UK over £2 billion annually [2]. In 2017 alone, nearly 65,000 people in England suffered a hip fracture [3]. Vertebral fractures of the thoracic and lumbar spine are often undiagnosed, despite being a common osteoporosis manifestation. When actively sought, these fractures account for nearly half of fragility fractures [4]. Systematic reviews indicate a ‘number needed to screen’ of 43 individuals aged 75 or older to prevent one vertebral fracture [5]. Opportunistic screening during routine radiology procedures, such as Computed Tomography (CT), could facilitate earlier diagnosis of bone fragility and reduce future fractures through timely treatment in a cost-effective manner [6], [7].

Despite National Osteoporosis Guidelines [8] advocating for early referral of patients with recent or multiple vertebral fractures, many individuals with osteoporosis continue to present late, often with height loss, chronic pain, mobility issues, and other complications [4]. Prescribing osteoporosis medication following reporting of vertebral fractures by radiologists is particularly poor [9]. Timely diagnosis and a growing array of effective treatments can significantly enhance the quality of life for osteoporosis patients and also reduce morbidity and mortality. Additionally, untreated spinal osteoporosis is progressive, with a fourfold increase in the risk of subsequent pathological vertebral fractures in the year following the first fracture [10].

Demand for CT diagnostics has surged, with scans performed in England rising from 1 million in 1996 to 5.67 million in 2018/19 [11]. Recent studies report 32-43% of over-60s undergoing CT scans for unrelated indications were also found to have osteoporosis or vertebral fractures [12], [13]. This indicates older CT attendees represent a high-risk population [13]. However, the diagnostic potential of CT for osteoporosis remains underutilised, with radiologists reporting only 16% of vertebral fractures in routine practice [14].

Several software packages exist that use routinely acquired CT images to diagnose osteoporosis in the spine and hip, as well as to identify and grade vertebral fractures. Notably, femoral neck bone mineral density (BMD) calculated from CT images using the Mindways Quantitative Computerised Tomography (QCT) software suite is a recognized alternative to standard Dual-energy X-ray Absorptiometry (DXA) BMD for inputting data into the fracture risk assessment tool, FRAX [15] [16]. We worked with Mindways QCT software developers to add a vertebral fracture identification Measurement Tool (MT), which we describe as SlicePick-MT [17].

At Cambridge University NHS Hospital, we have been designing and testing a ‘one-stop’ CT-based osteoporosis screening pathway. Approaching patients in CT waiting rooms about to undergo abdominal or pelvic CT scans, we invite older men and women (eligible for targeted case finding by age and sex criteria of NICE 146 osteoporosis guideline) to fill in a modified patient-completed version of the FRAX 10-year fracture risk estimation questionnaire. If FRAX indicates an elevated fracture risk, we digitally retrieve and reuse their clinical CT scans for bone density assessment and vertebral fracture identification. We currently do this with CliniQCT bone density software and SlicePick-MT (part of QCTPro software suite, Mindways Software Inc, TX, USA), which also usefully generates our semi-automated management advice for general practitioners (GPs). Beyond Cambridge, images from routine CT scans of patients attending any National Health Service (NHS) hospital can be easily transferred to the Cambridge hub via the National Image Exchange Portal, allowing us to measure BMD from patients at other hospitals, provided each CT scanner has been calibrated using the same ‘phantom’ device[17]. This allows remote diagnosis of osteoporosis, improved fracture risk calculation, and identification of fractured vertebrae [18].

By combining timely, opportunistic diagnosis of osteoporosis and/or vertebral fractures with a comprehensive individual bone health management plan sent to patient GPs, the service aims to improve patient care and health outcomes. This approach has the potential to enhance treatment rates and reduce incidence and burden of future fragility fractures. We report here on the study designed to test the feasibility of a subsequent definitive trial investigating the clinical effectiveness and cost-effectiveness of the screening service. We named our study PHOENIX-f (Picking up Hidden Osteoporosis Effectively during Normal CT Imaging without additional X-rays- feasibility).

## Methods

### Study design

This PHOENIX-f study was designed as a three-arm, non-blinded feasibility randomised controlled trial to test the feasibility of the CliniQCT pathway and its evaluation. The main outcomes were the ability to: i) randomise 375 ‘at risk’ participants within 10 months; ii) collect follow-up data on 75% or more of surviving participants at 1 year (specifically, completion of postal questionnaire items about receiving bone active medication). Secondary outcome measures included: i) prevalence of osteoporosis diagnosed with Mindways QCT and ii) prevalence of vertebral fractures identified with SlicePick-MT, both using CT scans acquired at enrolment; iii) self-reported use of bone active medication, among those with these identified conditions, at 1-year follow-up; and iv) survival. Technical and data flow aspects needed to run a definitive trial (intending to improve the treatment of osteoporosis across East Anglia) were also evaluated. A study protocol has been published [18].

### Enrolment and randomisation

Participants were recruited from four hospital sites around the East of England (Bedford, Bury St Edmunds, Huntingdon and Peterborough) plus the ‘hub’, Cambridge University Hospital NHS Trust. Participants were aged between 65-90 (females) or 75-90 (males) and attending hospital for an ordinary diagnostic or surveillance CT scan, where the spine or hips were visible in scan images, for clinical indications unrelated to bone health. Patients were excluded if: they had metalwork in their hips (given this creates a ‘streak’ artefact on the contralateral hip on scan images); were having a prone CT (such images cannot currently be processed by the Mindways software); or self-identified as already receiving prescription drugs for osteoporosis (such as bisphosphonates, denosumab, raloxifene, teriparatide or romosozumab).

Patient identification started in radiology CT waiting rooms, while waiting for a scan for an indication unrelated to bone health. The receptionist distributed patient invitation packs and self-consent forms; patients could request assistance with form completion. The novel patient invitation packs included a participant information sheet, pre-screening questions relating to previous hip surgery and prior or current use of bone active medications, a self-consent form, and the FRAX questionnaire required to calculate their initial FRAX 10-year fracture risk estimates (<u>data supplement file 1</u> of [18]). Patients in the ‘red’ (high) and ‘amber’ (moderate) risk categories – according to the UK National Osteoporosis Guideline Group (NOGG) FRAX fracture risk estimates – were subsequently randomised [using Research Electronic Data Capture (REDCap) software] to:

#### Group 1 – CliniQCT One-Stop vertebral fracture and osteoporosis pathway

The patient’s CT images already archived to the hospital’s Picture Archiving and Communication System (PACS) were sent to Cambridge and used for computer-aided diagnosis of vertebral fractures plus measurement of QCT bone density. This was performed by Cambridge NHS bone density technicians using CliniQCT and SlicePick-MT (Mindways QCT Pro) software, alongside regular quality control and CT phantom assurance procedures to overcome common sources of bias in volumetric BMD measurement [19]. Assessment results were sent in a semi-automatically generated [18] e-letter from the consultant in Cambridge to the patient’s GP. This includes: femoral neck and/or spine bone density results; vertebral fracture diagnosis specifying any level affected; adjusted 10-year risk of major osteoporotic fracture estimated from FRAX + QCT BMD; and recommended follow-up investigations and treatment text based on UK intervention thresholds in line with National Institute for Health and Clinical Excellence (NICE) guidelines [clinical decision tree, <u>data</u> <u>supplement file 2</u> of [18]].

#### Group 2 – FRAX Only (active control)

FRAX questionnaire responses were sent to patient GPs, who could then use the FRAX tool to assess the 10-year fracture risk and follow NOGG guidance. Delayed intervention with the same pathway as Group 1 was postponed until after one-year follow-up when ‘rescue analysis’ to identify vertebral fractures and diagnose osteoporosis followed the same procedures, including reports to GPs with treatment recommendations, as indicated, a year later.

#### Group 3 – Usual Care (passive control)

Patient GPs were informed by letter of their involvement in the study; no further information was shared then, beyond the usual CT scan report related to the original indicative condition. As with Group 2, ‘rescue analysis’ reports with treatment recommendations, as indicated, were sent to GPs after one-year follow-up.

### Data collection

Baseline and one-year follow-up postal questionnaires were sent to all randomised participants to collect data on quality of life and resource use relevant to bone health for health economic analysis. Mortality status was checked using NHS England Digital’s National Health Service Spine before re-contacting patients for follow-up, if not already flagged as deceased on hospital systems, and again before data were locked for analysis.

### Delayed intervention for control groups

To ensure ethical acceptability and promote patient safety, Group 1’s pathway (CliniQCT One-Stop and SlicePick) was applied for all surviving participants in Groups 2 (FRAX Only) and 3 (Usual Care) after follow-up. Thus, *all* moderate or high FRAX risk patient CT scans were analysed to calculate bone density and identify vertebral fractures, and their GPs were sent an e-letter containing diagnostic treatment and management advice, either at baseline or a year later. Further, one year after screening, CT scans of the non-randomised patients (since rated at low risk of fracture – FRAX ‘green’ scores) were also analysed by Mindways QCT, to ascertain the sensitivity and specificity of the FRAX screening process in CT-attending patients.

### Sample size considerations, randomisation and outcomes

Sample size considerations and outcomes to be measured in the feasibility trial have been previously reported in detail (7). In brief, the feasibility trial aimed to recruit and randomise 375 eligible participants uniformly across the three trial arms (i.e. 125 per arm) within 10 months. Key feasibility outcomes included ability to recruit to target, participant retention rates after 1 year follow-up and differences in osteoporosis treatment rates among survivors. Block randomisation was conducted online using REDCap stratified by study site (n=5), sex, and age group (female: <75 vs ≥75 years, male: <80 vs ≥80 years). Power calculations assumed type I error probability of 0.05 for the feasibility outcomes.

It was estimated that achieving the target sample size of 375 participants randomised would enable estimation of the trial’s response rate with a precision of 1.6% (half-width of the 95% confidence interval [CI]) assuming 40% of invited participants would be eligible (i.e. have red/amber FRAX score classification) and expecting a 30% response rate among those eligible.

For the 1-year retention outcome, it was estimated that the target sample size would provide 80% power to detect a difference of at least 6% or larger in the overall trial retention rates from a hypothesised 75% retention (as observed in the SCOOP trial [20], [21], [22], [23]. Additionally, the same sample size would provide >80% power to detect at least 12.2% absolute difference in retention rates between trial arms (or RR=1.16) based on two-sided two-sample test of proportion at 5% significance level.

For the 1-year treatment rates outcome, assuming treatment percentages at 1-year follow-up in the One-Stop CliniQCT & SlicePick-MT pathway versus FRAX Only versus Usual Care arms were 20.3% vs 15.5% vs 4.5% respectively – based on local data from prior intervention piloting – it was estimated the target sample size would provide >96% power to detect a linear trend in treatment proportions based on a two-sided test of binomial proportions at 5% statistical significance level, and 93% power for a secondary two-degrees of freedom test of between-group contrasts in comparison with usual care. When allowing for up to 25% attrition due to non-differential dropout or incompleteness of outcomes, the remaining sample size of 280 participants would have >84% power to detect the same magnitude of differences.

### Statistical analysis

Analyses and reporting followed <u>CONSORT-f</u> guidelines for randomised controlled trials, as extended for feasibility studies. Baseline characteristics were reported overall and by trial arms, summarised as mean and standard deviation (SD) for normally distributed continuous variables, median and interquartile range (IQR) for skewed continuous variables, and number and percentages for categorical variables.

Intention to treat analysis was conducted for the feasibility trial outcomes, with groups analysed as randomised to assess the overall effects of group assignment on the outcomes. Subgroup analysis was conducted for the outcome of receiving osteoporosis treatment at 1 year follow-up according to whether such treatment was indicated or not, regardless of whether treatment recommendation had been sent to the GP at baseline (for Group 1) or following rescue analysis that was conducted after the feasibility trial completion (for Group 2 and Group 3).

Overall response rate was calculated as percentage of participants who consented among all those invited. Post-consent trial eligibility rate was calculated as percentage of participants randomised (i.e. with red or amber FRAX) among those who consented. A logistic regression model – adjusted for site, sex, and age – calculated odds ratios comparing trial arms with Group 3 (usual care) as the reference. One site recruited fewer than ten patients, so their data were combined with the next nearest hospital’s data. The subgroup analysis model included interaction between trial arms and treatment recommendation to estimate subgroup-specific odds ratios and corresponding adjusted 1-year treatment outcome percentages.

Uncertainty around all statistical estimates were quantified with 95% confidence intervals, and p < 0.05 was used as guide for statistical significance. Sensitivity analyses addressed the uncertain effects of potential trial exclusion of small subgroups of patients (n=6 or n=14) for whom protocol violations could not be definitively adjudicated (prior bone active medication history unclear). Analyses were conducted in Stata version 14.

### Health economic analysis

We explored feasibility of collecting data to conduct an economic evaluation in a future definitive study. We calculated completion rates of: i) questions about resource use relating to bone health (this focus was chosen to reduce confounding in this older patient group presenting with an alternative primary condition – such as cancer); ii) the EuroQoL EQ-5D-5L (intended to be utilised to calculate quality adjusted life years (QALYs) in any future economic evaluation). Response patterns and participant feedback were also reviewed to suggest refinements to related data collection tools and methods.

To inform any future evaluation, we also conducted an exploratory within trial economic evaluation, in the form of a cost-utility analysis (CUA). This was from the perspective of NHS and personal social service input into bone health (motivated above). Resources were costed using standard reference sources such as 2023/24 National Cost Collection [22][21][23] and QALYs were calculated from the EuroQoL EQ-5D-5L responses collected at baseline and follow-up as recommended by Manca et al [24], with utilities valued using the algorithm method proposed by Hernández-Alava & Pudney [25]. The time horizon was 12 months. Where participants died, at follow-up they were treated as having no costs beyond intervention costs, and zero utility. Analysis used descriptive statistics to compare unadjusted mean (with 95% confidence interval) total costs and QALYs between arms.

### Patient and public involvement

Patients were involved in the development and conduct of the PHOENIX-f study. We recruited patient representatives to help shape and refine the study to ensure acceptability to patients undergoing CT scans for other medical reasons. The PPI group reviewed participant information sheets, consent forms, and recruitment materials to improve clarity, accessibility, and tone. They provided feedback on effectively communicating the aim of re-using existing CT images for osteoporosis screening and contributed to development of the QCT results and recommendation letters to general practitioners.

During the trial, PPI representatives continued to attend Trial Management Group meetings and advised on strategies to enhance participant engagement and follow-up. No patients were involved directly in data collection or analysis. The PHOENIX team is committed to ongoing engagement with PPI representatives, and we will do this by including PPI members as co-applicants in similar studies.

## Results

### Enrolment and randomisation

Figure 1 shows the CONSORT diagram of participant identification, recruitment and allocation. A total of 1,828 individuals were assessed as eligible, by age and scan type, for study invitation packs between mid-October 2019 and mid-August 2020, 740 and 1,088 respectively before and after UK COVID-19 pandemic lockdown restrictions began. Of these, 9% (156/1,828) were not invited due to missing their scan appointment or being missed in the department. Approximately half (51%, 858/1,672) of those invited declined to take part, 13% (219/1,672) had exclusion criteria (see Figure 1) and over a third of invited patients (595/1672, 36%) consented and met inclusion criteria. Among consented patients, 382 were classified as at moderate or high fracture risk on FRAX and randomised: thus, the recruitment outcome (375 randomised in 10 months) was achieved. Four patients were randomised in error and three patients withdrew shortly after randomisation due to illness, still leaving even distribution across trial groups: CliniQCT One-Stop intervention (128), FRAX only (active control, 124), and Usual Care (control, 123).

**Figure 1.**
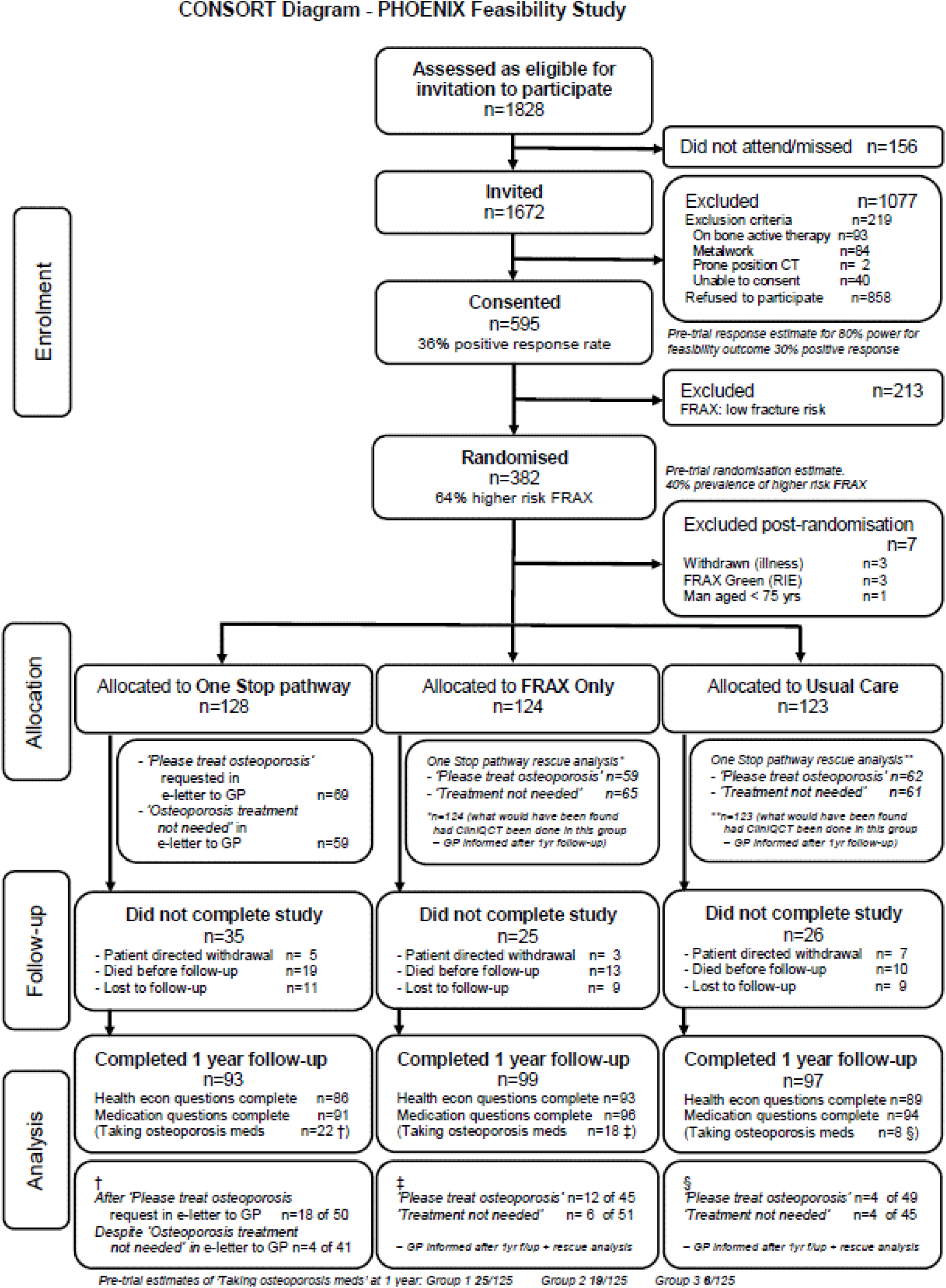
**CONSORT flow-chart: recruitment, randomisation and follow-up in the PHOENIX feasibility study**

### Mortality

During the follow-up year, 42 randomised patients died (see Figure 1); mortality rates were similar across the randomised sample study arms to mortality amongst non-randomised patients with lower fracture risk (green FRAX) scores: 42/375 (11%) and 27/213 (13%) respectively. Mortality differences between randomised groups were not statistically significant: CliniQCT One-Stop 19/128 (15%), Active Control 13/124 (10%), Usual Care 10/123 (8%); p=0.23.

### Retention

After one year across the three groups 289 of all 375 randomised patients (77%, 95% CI 73% to 81%) and 289 of 333 survivors at 1 year (87%, 95% CI 83% to 90%) were successfully followed-up: 93, 99 and 97 in the CliniQCT One-Stop Intervention, FRAX Only Active Control group and Usual Care groups respectively (see Figure 1). This co-primary outcome was therefore also met.

### Fracture risk at baseline

Table 1 summarises characteristics of the study sample. Randomisation achieved an even distribution across study groups of bone health related factors including age, sex and previous fragility fracture(s), and likewise similar prevalence of moderate or high FRAX fracture risk scores. Overall, 24% (89/375; 95% CI 19% to 28%) had fracture risk probability estimates classed as ‘high’ (red zone on FRAX risk plots).

**Table 1:** Characteristics of study participants at baseline – overall and by randomisation group.

|  | **Total**<br>*n*=375<br>n (%) or<br>median (IQR) | **One-Stop**<br>*n*=128<br>n (%) or<br>median (IQR) | **FRAX only**<br>*n*=124<br>n (%) or<br>median (IQR) | **Usual care**<br>*n*=123<br>n (%) or<br>median (IQR) |
| --- | --- | --- | --- | --- |
| **Female sex** | 335 (89%) | 113 (88%) | 112 (90%) | 110 (89%) |
| **Age at scan (years)** | 75 (70-80) | 75 (70-80) | 74 (71-81) | 75 (70-80) |
| **Age group** |  |  |  |  |
| 65-69 | 79 (21%) | 31 (24%) | 21 (17%) | 27 (22%) |
| 70-74 | 108 (29%) | 31 (24%) | 42 (34%) | 35 (28%) |
| 75-79 | 85 (23%) | 31 (24%) | 27 (22%) | 27 (22%) |
| 80-84 | 72 (19%) | 24 (19%) | 24 (19%) | 24 (20%) |
| 85+ | 31 (8%) | 11 (9%) | 10 (8%) | 10 (8%) |
| **Site** |  |  |  |  |
| Addenbrooke's | 255 (68%) | 84 (66%) | 85 (69%) | 86 (70%) |
| Peterborough | 33 (9%) | 11 (9%) | 12 (10%) | 10 (8%) |
| West Suffolk | 30 (8%) | 12 (9%) | 9 (7%) | 9 (7%) |
| Bedford | 57 (15%) | 21 (16%) | 18 (15%) | 18 (15%) |
| **Reason for CT** |  |  |  |  |
| Routine surveillance (known cancer) | 194 (52%) | 55 (43%) | 67 (54%) | 72 (59%) |
| Initial diagnostics (suspected cancer) | 69 (18%) | 25 (20%) | 25 (20%) | 19 (16%) |
| General medical diagnostics | 48 (13%) | 18 (14%) | 12 (10%) | 18 (15%) |
| Initial diagnostics (suspected benign pathology) | 15 (4%) | 8 (6%) | 3 (2%) | 4 (3%) |
| Surveillance (suspected benign pathology) | 10 (3%) | 3 (3%) | 6 (5%) | 1 (1%) |
| Unclear indication | 10 (3%) | 5 (4%) | 3 (2%) | 2 (2%) |
| Clinical trial | 6 (2%) | 2 (2%) | 1 (1%) | 3 (2%) |
| Information not available | 23 (6%) | 12 (9%) | 7 (6%) | 4 (3%) |
| **Prior fragility fracture** | 98 (26%) | 34 (27%) | 35 (28%) | 39 (24%) |
| **Initial fracture risk assessment** |  |  |  |  |
| FRAX estimated 10-year % risk of major osteoporotic fracture | 17 (13.0-23.0) | 17 (13.5-22.5) | 17 (13.0-22.5) | 17 (13.0-23.0) |
| FRAX estimated 10-year % risk of hip fracture | 7 (4.2-11.0) | 7 (4.7-11.5) | 6.3 (4.1-11.0) | 7.2 (3.9-11.0) |
| Moderate risk of fracture (FRAX estimate in NOGG amber zone) | 286 (76%) | 96 (75%) | 99 (80%) | 91 (74%) |
| High risk of fracture (FRAX estimate in NOGG red zone) | 89 (24%) | 32 (25%) | 25 (20%) | 32 (26%) |

Notes:
IQR Interquartile range
CT Computerised Tomography
NOGG National Osteoporosis Guideline Group

### Osteoporosis, vertebral fractures and recommendations for bone protection

CT scans were reviewed using QCT at baseline for the CliniQCT One-Stop pathway group and after 1-year follow-up for both control groups. Bone density measurements calculated by QCT and assessment for vertebral fractures informed treatment recommendations sent to GPs as indicated for all except 16 patients (3 lost to follow-up, 6 who withdrew from the study, 7 who died before GP reports were due). A further two patients’ CT images were inadequate for BMD calculation. Table 2 shows high prevalence of osteoporosis, diagnosed in 151 patients from 357 scans analysable by QCT for BMD calculation (42%, 95% CI 37% to 47%). Vertebral fractures were identified from 46 of the 258 scans which included the full thoraco-lumbar spine (18%, 95% CI 13% to 22%). Almost all patients’ CT scan imaging included at least one vertebrae, and of these 52/357 (15%, 95% CI 11% to 19%) were found to have at least one vertebral fracture. However, for those without evidence of fracture in their partial spine CT, it is not possible to confirm that their non-visualised vertebrae were fracture free. Amongst patients with any vertebral fracture prevalence of multiple vertebral fractures was high: 22/52 (42%, 95% CI 29% to 56%) had more than one. Over half of patients’ GPs (188/357, 53%, 95% CI 47% to 58%) were sent letters advising that osteoporosis treatment was recommended.

**Table 2:** Osteoporosis, vertebral fractures and treatment recommendation (assessments and recommendations to GPs made at baseline for One-Stop pathway patients and after one-year follow-up for patients randomised to FRAX Only and Usual Care trial arms)

|  | N | Total<br>mean (SD)<br>or n (%) | N | OneStop<br>pathway<br>mean (SD)<br>or n (%) | N | FRAX<br>Only<br>mean (SD)<br>or n (%) | N | Usual<br>Care<br>mean (SD)<br>or n (%) |
| --- | --- | --- | --- | --- | --- | --- | --- | --- |
| **Bone density measures from QCT** |  |  |  |  |  |  |  |  |
| Femoral neck BMD | 337 | 0.61 (0.12) | 119 | 0.60 (0.11) | 108 | 0.60 (0.12) | 110 | 0.61 (0.12) |
| Femoral neck T-Score | 337 | -1.7 (1.10) | 119 | -1.7 (1.00) | 108 | -1.7 (1.10) | 110 | -1.7 (1.10) |
| Total Hip T-Score | 174 | -2.0 (1.10) | 66 | -1.9 (1.10) | 57 | -2.0 (1.10) | 51 | -2.1 (1.20) |
| QCT lumbar vertebral BMD | 350 | 94.7 (31.7) | 127 | 92.6 (32.2) | 112 | 92.9 (30.8) | 111 | 98.9 (32.0) |
| **Diagnosed from QCT** |  |  |  |  |  |  |  |  |
| Osteoporosis at hip or spine | 357 | 151 (42%) | 128 | 55 (43%) | 113 | 47 (41%) | 115 | 49 (43%) |
| Vertebral fracture/s seen in CTs with complete thoracolumbar spine visible | 258 | 46 (18%) | 99 | 18 (18%) | 85 | 17 (20%) | 74 | 11 (15%) |
| Vertebral fracture/s seen in CTs with at least partial spine visible | 357 | 52 (15%) | 128 | 19 (15%) | 114 | 19 (17%) | 15 | 14 (12%) |
| >1 vertebral fracture seen in CTs with at least partial spine visible | 357 | 22 (6%) | 128 | 6 (5%) | 114 | 6 (5%) | 115 | 10 (9%) |
| **Bone protective treatment indicated and recommended** | 357 | 188 (53%) |  |  |  |  |  |  |
| GP sent recommendation to treat osteoporosis after baseline |  |  | 128 | 69 (54%) |  |  |  |  |
| GP sent recommendation to treat after follow-up 'rescue' analysis |  |  |  |  | 114 | 58 (51%) | 115 | 61 (53%) |

Notes:
N Number of observations in each analysis
SD Standard Deviation
QCT Quantitative Computerised Tomography analysis
BMD Bone Mineral Density

### Reported initiation of bone preserving treatment at one year follow-up

Table 3 describes the participants who completed the study with follow-up one year after randomisation. Comparison of Table 3 with Tables 1 and 2 show there was minimal change in the distribution of demographic and other bone health indicators between participants surviving to complete follow-up and baseline randomised cohort, confirming that the intervention was as applicable to survivors as to the baseline sample.

**Table 3:**
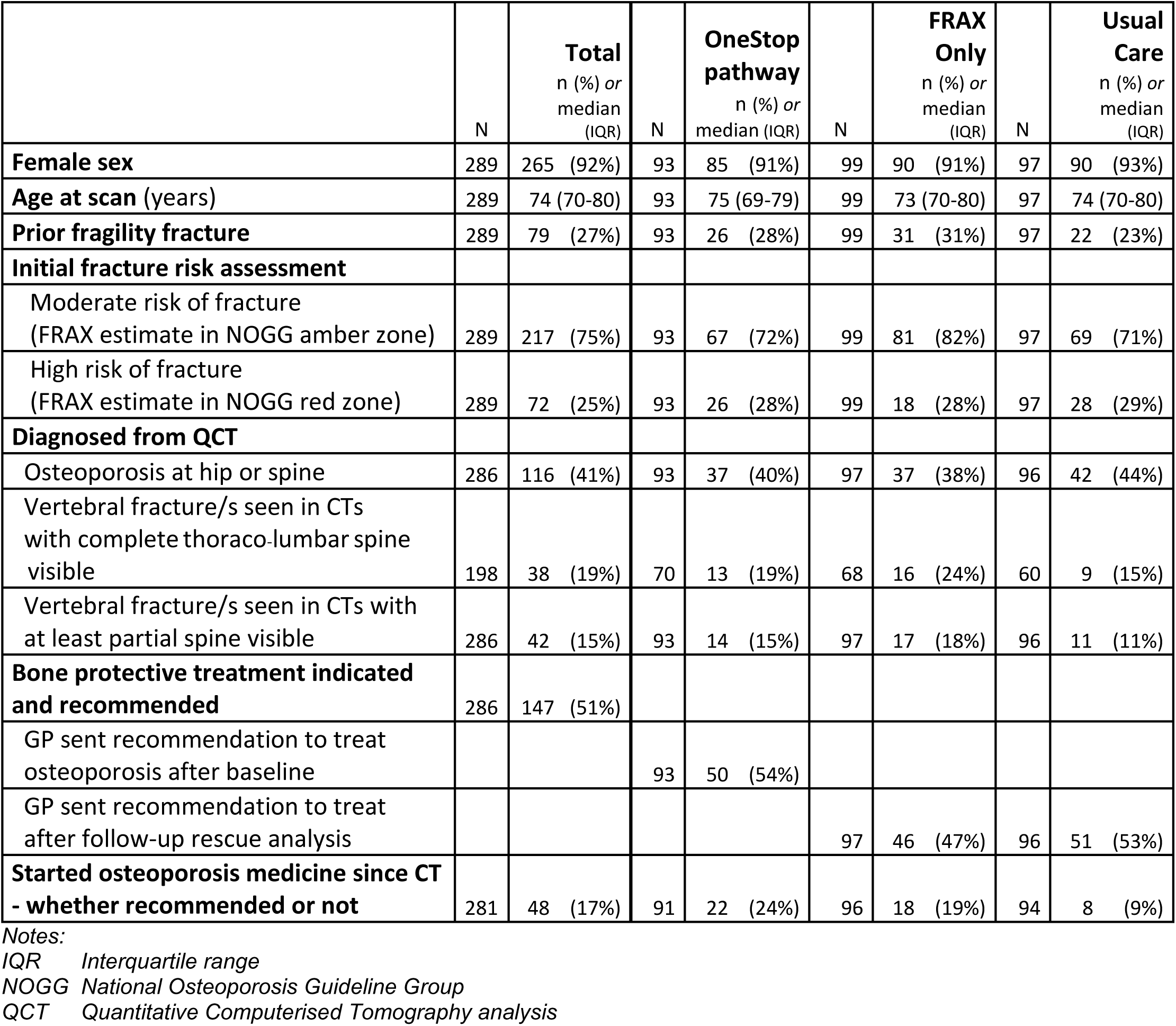
Fracture risk, osteoporosis, vertebral fractures and recommendation of bone preserving treatment for patients who completed one year follow-up.

|  | N | Total<br>n (%) or<br>median<br>(IQR) | N | OneStop<br>pathway<br>n (%) or<br>median (IQR) | N | FRAX<br>Only<br>n (%) or<br>median<br>(IQR) | N | Usual<br>Care<br>n (%) or<br>median<br>(IQR) |
| --- | --- | --- | --- | --- | --- | --- | --- | --- |
| **Female sex** | 289 | 265 (92%) | 93 | 85 (91%) | 99 | 90 (91%) | 97 | 90 (93%) |
| **Age at scan (years)** | 289 | 74 (70-80) | 93 | 75 (69-79) | 99 | 73 (70-80) | 97 | 74 (70-80) |
| **Prior fragility fracture** | 289 | 79 (27%) | 93 | 26 (28%) | 99 | 31 (31%) | 97 | 22 (23%) |
| **Initial fracture risk assessment** |  |  |  |  |  |  |  |  |
| Moderate risk of fracture<br>(FRAX estimate in NOGG amber zone) | 289 | 217 (75%) | 93 | 67 (72%) | 99 | 81 (82%) | 97 | 69 (71%) |
| High risk of fracture<br>(FRAX estimate in NOGG red zone) | 289 | 72 (25%) | 93 | 26 (28%) | 99 | 18 (28%) | 97 | 28 (29%) |
| **Diagnosed from QCT** |  |  |  |  |  |  |  |  |
| Osteoporosis at hip or spine | 286 | 116 (41%) | 93 | 37 (40%) | 97 | 37 (38%) | 96 | 42 (44%) |
| Vertebral fracture/s seen in CTs<br>with complete thoraco-lumbar spine<br>visible | 198 | 38 (19%) | 70 | 13 (19%) | 68 | 16 (24%) | 60 | 9 (15%) |
| Vertebral fracture/s seen in CTs with<br>at least partial spine visible | 286 | 42 (15%) | 93 | 14 (15%) | 97 | 17 (18%) | 96 | 11 (11%) |
| **Bone protective treatment indicated**<br>**and recommended** | 286 | 147 (51%) |  |  |  |  |  |  |
| GP sent recommendation to treat<br>osteoporosis after baseline |  |  | 93 | 50 (54%) |  |  |  |  |
| GP sent recommendation to treat<br>after follow-up rescue analysis |  |  |  |  | 97 | 46 (47%) | 96 | 51 (53%) |
| **Started osteoporosis medicine since CT**<br>**- whether recommended or not** | 281 | 48 (17%) | 91 | 22 (24%) | 96 | 18 (19%) | 94 | 8 (9%) |

Notes:
IQR Interquartile range
NOGG National Osteoporosis Guideline Group
QCT Quantitative Computerised Tomography analysis

Among patients who reported whether or not they were taking osteoporosis treatment at 1 year follow-up, differences between study groups in proportions with indication for treatment were non-significant [CliniQCT One-Stop 50/91 (55%); FRAX Only 45/96 (47%); Usual Care 49/94 (52%); p=0.53]. Of these patients, 18/50 (36%) reported taking bone active medication in the CliniQCT One-Stop arm, higher than in either of the other two groups: 12/45 (27%) in the FRAX Only Active Control group and only 4/49 patients (8%) in the Usual Care group (see Table 3). Adjusting for age, sex and recruiting hospital site minimally increased these proportions for the CliniQCT group (37%) and FRAX Only group (28%) (see Figure 2). Compared with receiving only usual care, patients who met treatment criteria were significantly more likely to report at follow-up that they were currently taking bone preserving treatment if in either the CliniQCT One-Stop Intervention (Odds Ratio 6.6, 95% CI 2.0 to 21.8, p=0.002) or Active Control groups (OR 4.9, 95% CI 1.4 to 16.8, p=0.01). However, confidence intervals around the proportions taking treatment in the two control groups overlapped (see Figure 2), as confirmed by the lack of significant difference in odds ratios between the CliniQCT One-Stop Intervention and the Active Control groups in terms of patients taking bone treatment (OR=1.4, 95% Cl 0.5 to 3.4, p=0.51). Sensitivity analyses tested effects of further exclusion of a further n=6 or n=14 patients with unclear history of possible prior bone active medication but found minimal changes in effect size.

**Figure 2:**
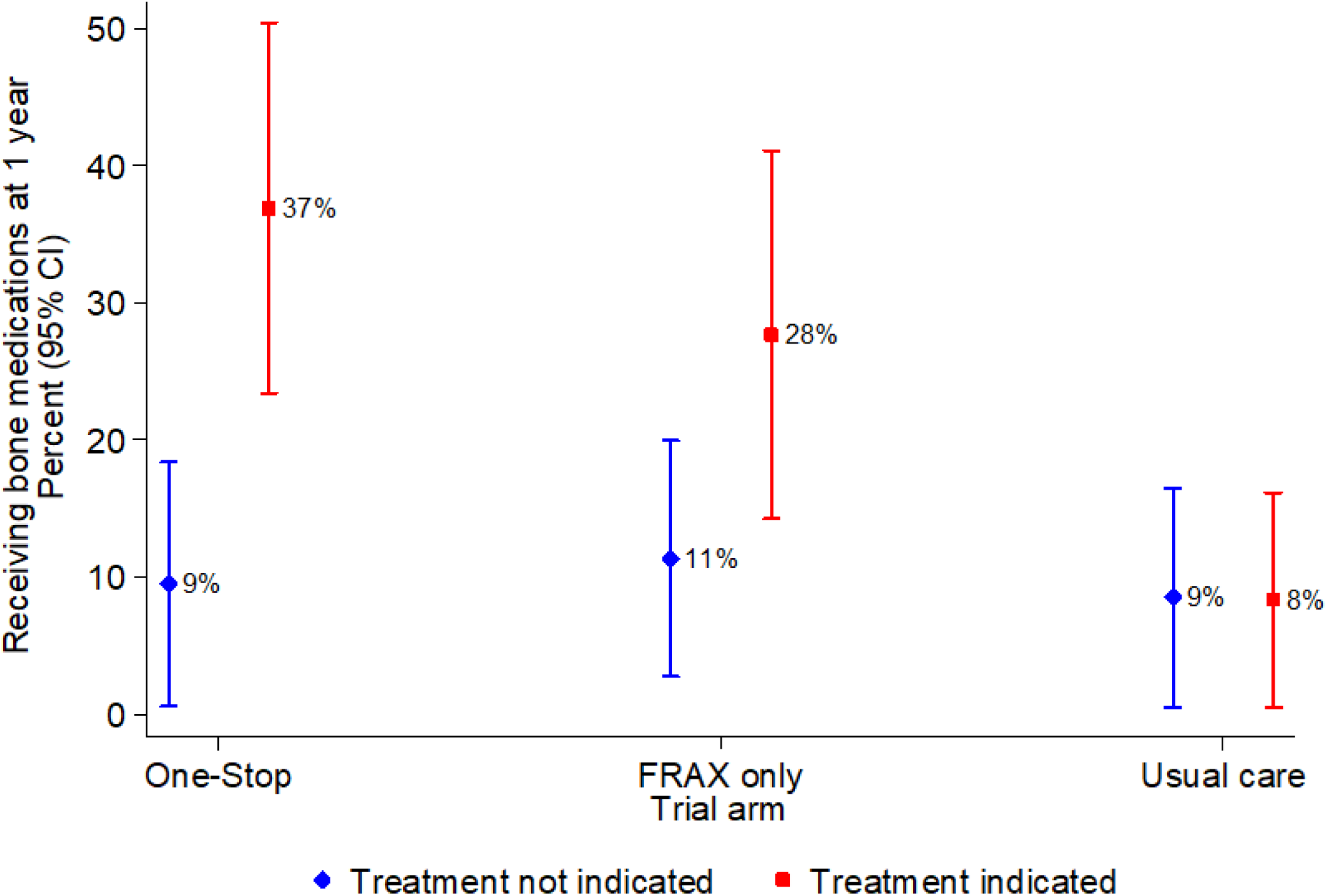
**Results of subgroup analysis on percentage of patients receiving bone medications at 1 year follow-up by trial arm according to whether there was guideline indication for bone medication treatment at baseline adjusted for site, sex and age**

Although bone preserving treatment was not indicated for nearly half of patients completing 1 year follow-up, approximately one in ten of these reported they were taking such treatments at follow-up, with minimal between groups differences: CliniQCT One-Stop pathway 4/41 (10%), FRAX Only 6/51 (12%), and 4/45 (9%) in Usual Care (see Figure 1). Age-, sex- and site-adjusted proportions were 9%, 11% and 9% respectively (see Figure 2).

Regardless of assessed treatment recommendations, comparing the three trial arms overall, at one year follow-up almost three times as many patients in the CliniQCT One-Stop intervention group, and more than twice as many patients in the FRAX Only Active Control group, reported taking osteoporosis medication, as compared with patients in the Usual Care group: 22/91 (24%), 18/96 (19%) and 8/94 (9%) respectively.

### Economic analysis

Completion rates on questions about use of NHS or personal social services related to bone health were high (>96%) for both time points, irrespective of arm (see Table S1). Rates of EQ-5D-5L completion were lower, but still high, particularly at follow-up (>83%; see Table S1). There were very few negative comments relating to completing either set of questions, though some patients highlighted challenges with completing the general health question of the EQ-5D-5L (item rating health on a visual analogue scale). Responses to resource use questions support refinement by having a separate item about scan use (e.g. MRI, CT, etc), and removing the social worker option, since no such contacts were reported.

Unit costs are reported in Table S2. We estimate the CliniQCT One-Stop and FRAX only interventions to cost £36.15 and £9.86 for each medium/high risk patient processed (these include a component for those low-risk patients who must be processed to identify higher risk patients). Exploratory economic evaluation results are given in Table 4. Usual Care ‘dominates’ the other options, having both lower total cost and accumulating higher QALYs; the CliniQCT One-Stop pathway dominates over FRAX Only. Costs are impacted by multiple outlier participants with high costs – illustrated in Figure S1 and with median costs primarily driven by the intervention costs (median total costs: CliniQCT One-Stop – £36.15; FRAX Only – £9.87; Usual Care - £0.00). These results should be interpreted with caution, given their exploratory nature.

**Table 4:** Exploratory economic evaluation results - mean [95% confidence intervals (CI)] total costs and QALYs, along with utility scores.

| **Mean (95% CI)** | **One-Stop pathway**<br>**(n=96)** | **FRAX Only**<br>**(n=100)** | **Usual Care**<br>**(n=94)** |
| --- | --- | --- | --- |
| **Total costs (£)** | 185 (6, 363) | 292 (118, 467) | 100 (-80, 178) |
| **QALYS** | 0.679 (0.634, 0.724) | 0.669 (0.625, 0.713) | 0.708 (0.662, 0.753) |
| **Baseline utility** | 0.738 (0.696, 0.779) | 0.699 (0.659, 0.740) | 0.758 (0.716, 0.800) |
| **Follow-up utility** | 0.621 (0.558, 0.683) | 0.639 (0.577, 0.700) | 0.657 (0.594, 0.721) |

## Discussion

We conducted a multi-centre, randomised, pragmatic feasibility trial of a one-stop service for older patients that screens for osteoporosis and vertebral fractures among those at higher risk, re-utilising CT scans taken for other purposes. If either are identified, the patient’s GP is informed with recommendations for bone preserving treatment. We confirmed the feasibility of recruitment (36% consent rate, 64% of whom randomised) and follow-up (76% retention, 86% of survivors) for a trial that compares this CliniQCT One-Stop pathway to both i) active control and ii) usual care. Secondary analyses also provided estimates of osteoporosis and vertebral fracture prevalence in this population and of this service’s effect on osteoporosis treatment rates – significantly higher among patients needing treatment in both the CliniQCT pathway and active control groups compared with usual care (36%, 26% and 8% respectively). Meeting these outcomes demonstrates the feasibility of rolling out this pathway model in a larger-scale effectiveness trial to evaluate its potential as a routine service. Given high return rates of postal questionnaires and high completion rates of resource use and utility questions, an economic evaluation in a future trial is likely feasible.

Despite the unexpected additional challenge of COVID-19 lockdown conditions during recruitment, we were still able to achieve our recruitment objective. Additionally, this afforded the opportunity to establish screening uptake when only delivered as an additional leaflet included within usual CT waiting room paperwork. We found that patients were broadly able to both complete the fracture risk questions (needed for identifying those at medium or high risk) and self-consent. High completion rates of the risk questions indicates the likely feasibility of the CliniQCT One-Stop pathway at this step, were it to be rolled out into routine practice, or indeed of only offering FRAX questionnaires.

We found clinical images were transferred easily from regional centres to the Cambridge central analysis hub (and critically could be ‘pulled’ in via NHS systems: this process was not reliant on being ‘pushed’ from each hospital), enabling us to re-use CT to measure bone density irrespective of where the scan was conducted.

Secondary outcome findings were surprising. Firstly, osteoporosis prevalence was higher than expected (42%). Vertebral fracture prevalence was also high (18% of 258 scans including the whole thoraco-lumbar spine) although within the range reported in this age-group [26], [27]. These reflect our finding that two-thirds of the patients consenting to fracture risk assessment had moderate or high risk of fracture – considerably higher than the 40% utilised in sample size calculations drawing from pilot data and prior published studies. This may be driven by the prioritisation/triaging to more acute patients necessary during COVID-19 – we might expect among them a higher prevalence of vertebral fractures and osteoporosis.

It was unexpected that the higher proportion of people receiving treatment in the CliniQCT One-Stop pathway at 1 year compared to usual care reached statistical significance in this trial powered for feasibility outcomes. Although encouraging that this effect was clear even in this relatively small trial, it is of concern that only approximately one in three treatment recommendations resulted in treatment at one year. It is unclear whether this is due to low prescription initiation by GPs or low uptake and adherence by patients – either is potentially affected by multiple factors that ongoing research seeks to understand. An ‘all secondary care’ approach – in which recommendations to treatment are acted upon by secondary care – might address some of these factors but could also be affected by service pressures.

Screening is an effective tool for reducing hip fractures: hip fracture incidence was reduced by 28% over five years using a screening approach based on FRAX and targeted DXA scanning [20]. The number needed to screen (NNS) found was 111 to prevent one hip fracture (National Institute of Health and Care Excellence, 2012). Hip fracture reduction relates to ‘at risk’ women receiving simple medication and was not thought to be due to altered health behaviours [29][30]. Whether screening for and/or diagnosing osteoporosis caused women unwarranted anxiety is unclear [31][32]. Our patients (both consenting and non-consenting) were asked for feedback on study procedures and on screening via their consent forms. A brief review of this feedback suggests that those patients who wanted to know about their bone health were very supportive and pleased to have this ‘added extra’ to their diagnostic or surveillance CT scan.

Almost half the loss to follow-up at one year was due to death, raising questions for future research and service design regarding best approaches to identifying high risk patients with life expectancy adequate to warrant starting bone protection.

Service provision factors relevant to bone health management likely affected patients in each arm equally despite some between hospital differences. At Cambridge University Hospital, radiologists routinely include an agreed code in all CT scan reports confirming whether a new or stable vertebral fracture, or none, is visible; however, this was not standard practice in the other hospitals from which study patients were recruited. During this study’s recruitment and follow-up period, there was no pathway across the study’s five hospitals for referring patients with newly identified vertebral fractures directly to the local Fracture Liaison Service or to any other specialist.

Since our study, Woisteschlager et al. demonstrated that using intravenous contrast leads to overestimation of spine vBMD [33]. We routinely include the following caveat when reporting contrast-enhanced spine vBMD: ‘*note Contrast-enhanced scan, may overestimate BMD’.* However, Woisteschlager et al. usefully confirm any patient with spine vBMD values below 80mg/cm3 will have been correctly identified as below a fragility threshold; however, absolute BMD could be up to 16% lower. Consequently, including contrast-enhanced spine QCT may result in some treatment eligible patients going untreated.

Overall, this study showed that delivery of this screening pathway and our research methods to evaluate its effectiveness are both feasible. Patients could consent themselves and provide the necessary demographic and clinical risk factor data. Older CT-attenders had high levels of bone health need, including vertebral fractures and osteoporosis, which were easily identified by FRAX and re-using CT scans. Providing bespoke and accurate diagnostic and treatment advice was simple and led to more than four times the number of patients receiving fracture-preventing therapy than usual care. Just alerting GPs to their patient’s FRAX risk factors was the pathway for our ‘passive control group’ in which treatment rates were also significantly higher than in the ‘usual care’ group but lower than in the CliniQCT One-Stop group, though the sample was too small for this difference to be statistically significant. To provide robust clinical- and cost-effectiveness data, the evidence base needs a larger trial powered to evaluate how these approaches compare and to further investigate whether treatment rates.

## Data availability

Data are available on reasonable request. Applications for access to the final trial dataset will be through the Trial Management Group (TMG) following a controlled access model (openly available to all applicants) as set out in the MRC and NIHR guidance (https://www.methodologyhubs.mrc.ac.uk/files/7114/3682/3831/Datasharingguidance2015.pdf).

## Transparency

The lead author (the manuscript’s guarantor) affirms that the manuscript is an honest, accurate, and transparent account of the study being reported; that no important aspects of the study have been omitted; and that any discrepancies from the study as planned (and, if relevant, registered) have been explained.

## Author contributions

KP is the lead investigator who conceived the study and secured NIHR funding jointly with EMC, JF, SK, LS, TT and APW, who all contributed to the study design. DC, JB, KP and KW conducted the trial, with initially KW and then DC coordinating study site recruitment and follow-up. JB and DC carried out all QCT analyses. Vertebral fracture review was by KP and TT. SK designed and carried out statistical analyses of the trial. APW designed the health economic analyses, carried out by HR and APW. KP and RS wrote the first draft of this paper, JF and APW contributed substantially to subsequent drafts, and all authors have reviewed, commented and accept the final version. The corresponding author attests that all listed authors meet authorship criteria and that no others meeting the criteria have been omitted.

## Ethics

National Research Ethics Service approval from East of England committee: REF/19/EE/0176

## Provenance and peer review

Not commissioned; externally peer reviewed.

## Acknowledgements

We acknowledge the essential contributions of our PPIE colleagues, Jeremy Dearling and Jane Marjoram, and of all the PHOENIX-f study participants, without which this research could not have been carried out, the assistance of the Eastern Research Design Service, the Cambridge Clinical Trials Unit (Prof Ian Wilkinson), the NIHR Portfolio and all those NHS research, clinical and administrative/reception staff and participants from Bedford, Bury St Edmunds, Huntingdon, Peterborough, and Addenbrooke’s hospitals. The Mindways software team of Wolfram Timm and J Keenan Brown. Information Governance at CUHNFT, the Sectra Imaging Exchange Portal team. Helen Adderley, CUHNFT radiology and the UKAS accreditation team.

