## Supplementary material for "PHOENIX-f: A Randomised Controlled Feasibility Study of One-Stop Osteoporosis Screening among Patients Undergoing Routine Computed Tomography using FRAX and CliniQCT Bone Density Measurement and Vertebral Fracture Detection": Table S1

**Table S1:**
**Completion rates of health economic forms and participants numbers available for complete case analysis**

| | | One-Stop pathway | FRAX Only | Usual Care | Overall |
|---|---|---|---|---|---|
| Baseline | Participants | 128 | 124 | 123 | 375 |
| | Complete* EQ-5D-5L | 84% | 88% | 87% | 86% |
| Follow-up | Participants followed-up | 93 | 99 | 97 | 289 |
| | Complete resource use | 97% | 98% | 96% | 97% |
| | Complete* EQ-5D-5L | 95% | 96% | 96% | 96% |
| Participants with complete data for economic evaluation (including those identified as dead before follow-up) | | 96 | 100 | 94 | 290 |

*'Complete' is defined as those individuals completing the five items of the EQ-5D-5L needed to calculate utilities.
