## Supplementary material for "PHOENIX-f: A Randomised Controlled Feasibility Study of One-Stop Osteoporosis Screening among Patients Undergoing Routine Computed Tomography using FRAX and CliniQCT Bone Density Measurement and Vertebral Fracture Detection": Table S2

**Table S2:**
**Unit costs (£, 2023/2024) used in the economic evaluation** [21], [22], [23]

| Cost component | Cost £, 2023/24 | Source, detail and assumptions |
|---|---|---|
| **Identification of higher risk (FRAX red/amber) patients (cost per patient)** | **5.70** | |
| Identifying CT list patients having spine or hips scanned & meeting age criteria | 3.55 | Assume 30 minutes per working day of a Agenda for Change Band 3 Administrator [PSSRU, Table 12.2.4], annual salary £22,562. Assume 48 working week year, 37.5 hours per week, to recruit 382 higher risk patients over ten months (implicitly includes identifying those patients that turn out to be lower risk/FRAX green) |
| Scanning & emailing FRAX questions for review | 1.87 | Assume 5 minutes per patient of an Agenda for Change Band 3 Administrator [PSSRU, Table 12.2.4], annual salary £22,562. Assume 48 week working weeks, 37.5 hour per week. Inflate to take account of also processing lower risk/FRAX green in the trial ratio (382:213) |
| Printing bone health questionnaire | 0.14 | Printing an A4 side at 0.07665 (1000 pages@£76.65 from A4 Flyers & Leaflets Printing, Print A4 Leaflets Online UK \| instantprint). Inflate to take account of also processing lower risk/FRAX green in the trial ratio (382:213) |
| Printing FRAX questionnaire | 0.14 | Printing an A4 side at 0.07665 (1000 pages@£76.65 from A4 Flyers & Leaflets Printing, Print A4 Leaflets Online UK \| instantprint). Inflate to take account of also processing lower risk/FRAX green in the trial ratio (382:213) |
| **CliniQCT One-Stop assessment and dissemination to GP (cost per processed patient)** | **30.45** | |
| Calculation of 10-year FRAX risk score (red, amber, green) | 5.83 | Assume 5 minutes per patient of an Agenda for Change Band 5 hospital-based scientific and professional staff [PSSRU, Table 11.1.2], £39 per working hour. Inflate to take account of also processing lower risk/FRAX green in the trial ratio (382:213) |
| Bone health assessment | 11.38 | Assume 17.5 (15-20) minutes per patient of an Agenda for Change Band 5 hospital-based scientific and professional staff [PSSRU, Table 11.1.2], £39 per working hour. Assessment includes: CT scan review; cleaning CT data; Mindways QCT Pro analysis; bone density (CT-BMD) calculated; and vertebral fractures identified. |
| Clinician-verified report | 9.08 | Assume 5 minutes per patient of a medical consultant [PSSRU, Table 11.3.2], £109 per working hour. Report approval and treatment advice. |
| Processing FRAX/Mindways report for posting to GP | 3.25 | Assume 5 minutes per patient of an Agenda for Change Band 3 Administrator [PSSRU, Table 12.2.4], annual salary £22,562. Assume 48 week working weeks, 37.5 hour per week. Inflate to take account of also processing lower risk/FRAX green in the trial ratio (382:213) |
| Printing FRAX/Mindways report for GP | 0.15 | Printing 2xA4 sides at 0.07665 (1000 pages@£76.65 from A4 Flyers & Leaflets Printing, Print A4 Leaflets Online UK \| instantprint). |
| Postage costs of report for GP | 0.76 | Second class stamp, Royal Mail. |
| **Active control intervention (FRAX only; cost per processed patient)** | **4.16** | |
| Processing FRAX/Mindways report for posting to GP | 3.25 | Assume 5 minutes per patient of an Agenda for Change Band 3 Administrator [PSSRU, Table 12.2.4], annual salary £22,562. Assume 48 week working weeks, 37.5 hour per week. Inflate to take account of also processing lower risk/FRAX green in the trial ratio (382:213) |
| Printing FRAX/Mindways report for GP | 0.15 | Printing 2xA4 sides at 0.07665 (1000 pages@£76.65 from A4 Flyers & Leaflets Printing, Print A4 Leaflets Online UK \| instantprint). |
| Postage costs of report for GP | 0.76 | Second class stamp, Royal Mail. |

| Cost component | Cost £, 2023/24 | Source, detail and assumptions |
|---|---|---|
| **Hospital admissions (cost per admission)** | | |
| Day case (musculoskeletal) | 1700.00 | Weighted average of national average unit cost for musculoskeletal system day case [NCC] |
| A&E attendance | 273.00 | Weighted average of national average unit cost of Emergency care [NCC] |
| Non-elective inpatient short stays (musculoskeletal) | 988.00 | Weighted average of national average unit cost for musculoskeletal non-elective inpatient short stays, admitted patient care [NCC] |
| Non-elective inpatient long stays (musculoskeletal) | 7613.00 | Weighted average of national average unit cost for musculoskeletal non-elective inpatient long stays, admitted patient care [NCC] |
| Hip replacement | 8358.00 | Most common elective, non-trauma hip replacement activity [NCC] |
| **Outpatient visits and procedures (cost per procedure/appointment)** | | |
| Consultant rheumatologist (face to face, follow-up) | 194.00 | Consultant led Rheumatology Service, Non-Admitted Face-to-Face Attendance, Follow-up [NCC] |
| Hospital nurse | 22.68 | Band 5, hospital nurse [PSSRU, Table 11.2.2]. Cost per working hour £42. Assume 54% of time spent with patient [PSSRU, table 12.5] |
| CT scan | 103.00 | Computerised Tomography Scan of One Area, without Contrast, 19 years and over, directly accessed diagnostic services [NCC] |
| X-ray | 42.00 | Plain film, directly accessed diagnostic services [NCC] |
| Nuclear scan | 277.00 | Cost of Dex scan £81 plus administration cost £196. Assumed administration cost covered by HRG code WF01B, chosen most common (Trauma and Orthopaedic Service) £196 [NCC] |
| MRI | 165.00 | Magnetic Resonance Imaging Scan of One Area, without Contrast, 19 years and over, directly accessed diagnostic services [NCC] |
| DXA | 81.00 | Dual-energy X-ray Absorptiometry scan, directly accessed diagnostic services [NCC] |
| **Primary and community care visits (cost per visit)** | | |
| GP | 49.33 | Costs for General Practitioner (GP) [PSSRU, p.63]. £49 per surgery consultation lasting 10 minutes. Assume 10 minute face-to-face consultation |
| Practice nurse | 7.83 | Costs for GP practice nurse [PSSRU, p.62]. Agenda for change band 5. Unit cost per hour (without qualifications). Assume 10 minute face-to-face consultation |
| Community physiotherapist | 78.00 | Community services, average cost per one-to-one session for physiotherapy £78 [PSSRU, p.36] |
| **Pharmaceuticals (monthly cost)** | | |
| Alendronic acid 70mg tablets | 2.48 | 70mg taken once weekly. £2.48, 4 per item [PCA] |
| Risedronate sodium 35mg tablets | 2.21 | 35mg taken once weekly. £2.21, 4 per item [PCA] |
| Ibandronic acid 150mg tablets | 2.25 | 150mg taken once monthly. £2.25, 1 per item [PCA] |
| Raloxifene 60mg tablets | 5.21 | 60mg once daily, £4.86 per item, 28 per item [PCA] |
| **Pharmaceuticals requiring administation (per dose)** | | |
| Denosumab injection | 207.43 | Assume administered at GP surgery once every 6 months. Drug cost £183.93 per dose [PCA]. Assume GP nurse 30mins [PSSRU] |
| Zoledronate infusion | 370.14 | Assume IV administered once per year. Drug cost £174.14 [PCA]. Assumed administration cost covered by HRG code WF01B, chosen most common (Trauma and Orthopaedic Service) £196 [NCC]. Where infusion listed but no drug name given, used Zoledronate as most commonly prescribed (https://www.mkuh.nhs.uk/wp-content/uploads/2019/03/Response-4342.pdf) |

| Cost component | Cost £, 2023/24 | Source, detail and assumptions |
|---|---|---|
| Pamidronate infusion | 306.00 | Assume IV administered and once per 6 months. Drug cost £110 [PCA]. Assumed administration cost covered by HRG code WF01B, chosen most common (Trauma and Orthopaedic Service) £196 [NCC] |
| **Additional pharmaceuticals (monthly cost)** | | |
| Colecalciferol 400unit / Calcium carbonate 1.5g chewable tablets | 3.54 | 400unit dose. £3.54 per item, 30 per item [PCA] |
| Adcal-D3 chewable tablets tutti frutti | 4.02 | Two per day. £3.75 per item, 56 per item [PCA] |
| Alfacalcidol 250nanogram capsules | 6.34 | Maintenance 0.25–1 microgram daily. £6.34 per item, 30 per item [PCA] |
| Calceos 500mg/400unit chewable tablets | 4.01 | Two per day. £4.01 per item, 60 per item [PCA] |
| Evacal D3 1500mg/400unit chewable tablets | 2.98 | Two per day. 2.78 per item, 56 per item [PCA] |
| TheiCal-D3 1000mg/880unit chewable tablets | 3.15 | One per day. £3.15 per item, 30 per item [PCA] |
| Iasibon 50mg tablets | 209.73 | One per day. £195.75 per item, 28 per item [PCA] |
| Accrete D3 One a Day 1000mg/880unit chewable tablets | 3.19 | £3.19 per item, 30 per item [PCA] |
| Utrogestan 100mg capsules | 13.09 | £15.71 per item, 30 per item. 1 capsule (100mg) once a day, on days 1 to 25 of 28-day HRT cycle [PCA] |
