## Supplementary figures and images for "PHOENIX-f: A Randomised Controlled Feasibility Study of One-Stop Osteoporosis Screening among Patients Undergoing Routine Computed Tomography using FRAX and CliniQCT Bone Density Measurement and Vertebral Fracture Detection"

### Figure S1

**Figure S1:**
**Estimated total costs, split by arm, in the exploratory economic evaluation**

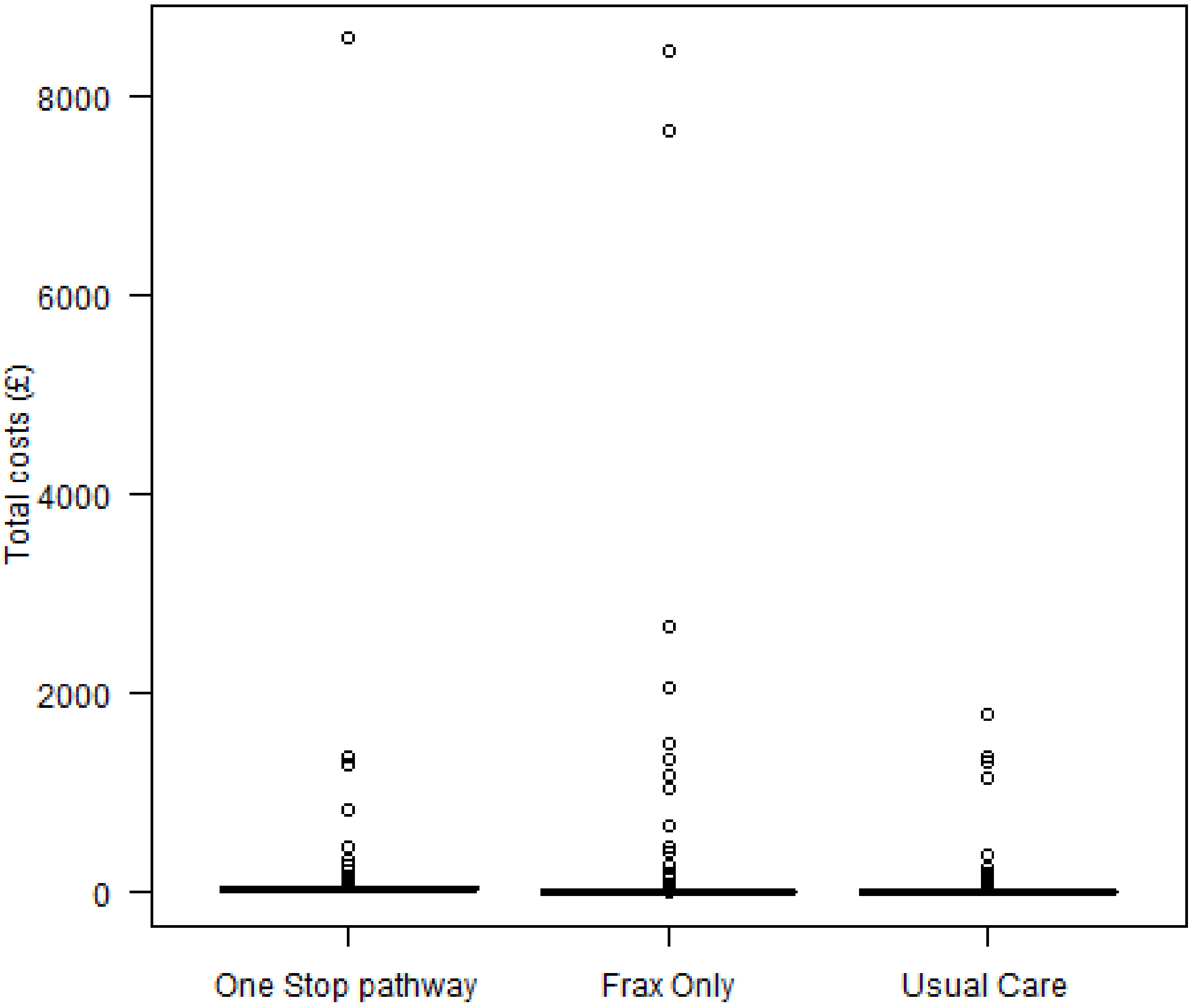
